# IMPACT OF COVID-19 PANDEMIC ON SICKNESS ABSENCE FOR MENTAL ILL HEALTH IN NATIONAL HEALTH SERVICE STAFF

**DOI:** 10.1101/2021.06.09.21258629

**Authors:** Diana A van der Plaat, Rhiannon Edge, David Coggon, Martie van Tongeren, Rupert Muiry, Vaughan Parsons, Paul Cullinan, Ira Madan

**Affiliations:** National Heart and Lung Institute (NHLI), Imperial College London, UK; Lancaster University, Lancaster Medical School, Bailrigg, Lancaster, UK; MRC Lifecourse Epidemiology Unit, University of Southampton, UK; University of Manchester, Centre for Occupational and Environmental Health, School of Health Sciences, Manchester, UK; Occupational Health Service, Guy’s and St Thomas’ NHS Foundation Trust, UK; King’s College London Faculty of Life Sciences and Medicine, London, UK

**Keywords:** Mental ill health, Sickness absence, healthcare workers, COVID-19

## Abstract

**Objective:** To explore the patterns of sickness absence in National Health Service (NHS) staff attributable to mental ill health during the first wave of the Covid-19 epidemic in March – July 2020

**Design:** Case-referent analysis of a secondary data set

**Setting:** NHS Trusts in England

**Participants:** Pseudonymised data on 959,356 employees who were continuously employed by NHS trusts during 1 January 2019 to 31 July 2020

**Main Outcome Measures:** Trends in the burden of sickness absence due to mental ill health from 2019 to 2020 according to demographic, regional and occupational characteristics.

**Results:** Over the study period, 164,202 new sickness absence episodes for mental ill health were recorded in 12.5% (119,525) of the study sample. There was a spike of sickness absence for mental ill health in March-April 2020 (899,730 days lost) compared with 519,807 days in March-April 2019; the surge was driven by an increase in new episodes of long-term absence and had diminished by May/June 2020. The increase was greatest in those aged >60 years (227%) and among employees of Asian and Black ethnic origin (109%-136%). Among doctors and dentists the number of days absent declined by 12.7%. The biggest increase was in London (122%) and the smallest in the East Midlands (43.7%); the variation between regions reflected the rates of Covid-19 sickness absence during the same period.

**Conclusion:** Although the Covid-19 epidemic led to an increase in sickness absence attributed to mental ill health in NHS staff, this had substantially declined by May/June 2020, corresponding with the decrease in pressures at work as the first wave of the epidemic subsided.

**Article Summary:** **Strengths and limitations of this study**

- Large study population
- Study population were not self-selected
- Job exposure matrix allowed adjustment for occupational exposure
- Data did not extend to the start of the second wave in September 2020

## Background

The National Health Service (NHS) is the largest employer in England, with almost a million staff. As in the wider national workforce, mental ill health is a major cause of sickness absence among NHS employees (Copeland, 2019; ONS, 2021). We have previously highlighted a clear increase in such absence during the first wave of the Covid-19 pandemic (March to July 2020) as compared with the corresponding period in 2019 (Edge et al., 2021). The trend contrasted with that for other disorders such as cancer, gastrointestinal and gynaecological disease, musculoskeletal complaints, and injuries, for all of which, rates of new sickness absence declined.

The rise in mental ill health may have resulted from increased stress, at or away from work, as a consequence of the pandemic. However, it could also reflect longer term trends that began before Covid-19 emerged. Better understanding is needed, both as a pointer to possible preventive strategies, and also to inform the optimal deployment of personnel when healthcare services are under severe pressure.

We therefore undertook a more detailed exploration of patterns of sickness absence for mental ill health in NHS staff between January 2019 and June 2020. Specifically, we were interested in whether there was a step-change in new absences for mental ill health when the Covid-19 epidemic began, whether trends differed for long-term and shorter episodes of absence, and whether they applied differentially to particular demographic and staff groups.

## Methods

With approval from the NHS Health Research Authority (reference 20/SC/0282), we were granted access to pseudonymised data that had been abstracted from the NHS Electronic Staff Record (ESR) on all individuals who had been continuously employed by NHS trusts in England from 1 January 2019 to 31 July 2020 (the study period). Details of the information obtained, and its preliminary processing have been reported in a supplement to an earlier paper (van der Plaat et al., 2021). For each member of staff, the data included demographic and occupational characteristics, and the start and end dates of all episodes of sickness absence during the study period (other than for annual leave) with the reason for absence.

For this paper, we focused principally on sickness absence for mental ill health, but to check on the specificity of some findings, we also examined absences for back problems and for other musculoskeletal disorders which are common in this population. Each category of absence was identified by a code that trusts use when entering records onto the centralised ESR database. Other variables that we analysed were: sex; age group at 15 September 2020 (nine categories); ethnicity (seven categories); staff group (10 categories), the region of the employing trust (nine categories), and Covid-19 sickness absence during March-April 2020. Where an individual had changed jobs during the study period, we defined staff group according to that which applied at the beginning of the period; the coding scheme for staff group was that used in the ESR database (NHS Digital, 2020). As in earlier reports (Edge et al., 2021; van der Plaat et al., 2021), Covid-19 sickness absence was defined as sickness absence in any of five categories (cough/flu, chest/respiratory, infectious diseases, other, unknown), for which Covid-19 was recorded as a related reason.

Statistical analysis was with R (version 4.0.4) software (Team, 2020). We first used logistic regression to explore risk factors for cumulative prevalence of new sickness absence because of mental ill health during 1 January 2019 to 30 June 2020, according to whether or not at least one episode continued for >28 days. Associations were summarised by odds ratios (ORs) with 95% confidence intervals (CIs).

To assess trends in the burden of sickness absence for mental ill health over the course of the study period, we then plotted three measures (total days of absence, number of new episodes of absence with duration ≤28 days, and number of new episodes with duration >28 days) for consecutive two-month intervals.

We next examined changes from 2019 to 2020 in total days lost through sickness absence for mental ill health during March and April, according to demographic and occupational characteristics. Finally, we explored the correlation across regions between the year-on-year change in days lost because of mental ill health in March and April and cumulative prevalence of Covid-19 sickness absence in March-April 2020.

As sensitivity analyses, we repeated the analyses excluding individuals in whom one or more of sex, age, ethnicity or the end date of a period of sickness absence for mental ill health was imputed because of inconsistencies, or whose job changed over the study period.

## Results

After exclusion of 21,775 individuals who were absent from work continuously throughout the study period analysis was based on 959,356 employees (77.0% female). Over the 18-month study period, a total of 164,202 new sickness absence episodes for mental ill health were recorded in 119,525 individuals (12.5% of the study sample). In combination with episodes that were already ongoing at 1 January 2019, these accounted for 6,255,602 days of absence, equating to between 1 and 2 percent of contracted time.

Although most absence for mental ill health was of short duration (≤28 days), almost 6% of employees experienced one or more longer episodes over the course of the study period. Table 1 shows the cumulative prevalence of new sickness absence for mental ill health in relation to various demographic and occupational characteristics. After adjustment for other factors, cumulative prevalence of absence, whether of short- or long-duration, was some 40% higher in women than in men. Prevalence of long-duration absence was highest in the older age groups (>30 years), whereas that of absence which was only ever of short duration, declined progressively across the age bands (OR 0.45 for age >60 vs. <25 years). Employees of non-white ethnicity tended to have lower cumulative prevalence, with ORs for Asian and Black relative to White workers ranging from 0.44 to 0.71. Also, there were notable differences by staff group, with the highest rates in ‘additional clinical services’ (a group that included care assistants) and much lower rates in healthcare scientists and medical/dental personnel. Relative to administrative and clerical workers, these three groups had adjusted ORs of 1.58, 0.54 and 0.32 respectively for long-duration absence, and 1.55, 0.73 and 0.26 for absence that was only ever of short duration.

After allowance for other characteristics, cumulative prevalence varied markedly by region, with the lowest rates in London and the South East, and the highest in the North East and North West. This applied particularly to long-duration absence (ORs relative to London 2.34 for North East and 2.36 for North West). These large regional differences were apparent also when analysis was restricted to specific staff groups. For example, in doctors and dentists, the adjusted OR for long-duration absence relative to London was 1.97 (95%CI 1.44-2.71) in the North East and 2.76 (95%CI 2.25-3.37) in the North-West.

To check on the specificity of these findings for mental ill health, we carried out a similar analysis for risk of long-duration (>28 days) sickness absence because of back problems and other musculoskeletal disorders (Supplementary Table S1). Risk relative to administrative and clerical workers was substantially elevated in additional clinical services (OR 2.57) and in estates and ancillary workers (OR 2.41), but again was low in healthcare scientists (OR 0.62) and doctors and dentists (OR 0.34). However, after adjustment for other risk factors, regional differences were smaller for long-term back problems and other musculoskeletal disorders than for long-term mental ill health. Risk was lowest in London and the South West, and highest in the North East and North West (ORs relative to London 1.59 and 1.53).

Figure 1 illustrates trends over the course of the study period in sickness absence for mental ill health. Total days of absence per two-month period increased over the first 10 months of 2019, and then declined somewhat, but with a clear spike in March and April 2020 (Figure 1A). The number of days lost in those two months (899,730) was substantially higher than in the corresponding period 12 months earlier (519,807). In contrast, fewer days were lost in May and June 2020 than in May and June 2019 (516,890 vs. 572,401). Likewise, total days of absence and numbers of new absences of short duration increased progressively during January to October 2019, but they then plateaued, with no marked increase in any of the subsequent two-month intervals (Figure 1B). The surge in total days of absence in March and April 2020 was driven by an increase in new episodes of long-duration absence (10,376 new episodes compared with 5,151 in March and April 2019). New episodes of long-duration absence were also more frequent in May and June 2020 than in the corresponding period a year earlier, although to a lesser extent (7,835 vs. 5,833) (Figure 1C).

Table 2 presents changes between 2019 and 2020 in the total days lost through sickness absence for mental ill health during March and April, with results shown separately for different demographic and occupational groups. A clear increase was apparent in almost all groups, but it was greater in women than in men (78.0% vs. 47.7%); in those aged <35 years (102% to 124%) and >60 years (227%); and in employees of Asian and Black ethnicity (109% to 136%). In contrast, the increase was smaller than average in registered nurses and midwives (53.8%), while among doctors and dentists, the number of days absent declined by 12.7%. Across the regions, the biggest increase was in London (122%), and the smallest increases were in the East Midlands (43.7%) and South West (45.8%).

Figure 2 plots percentage change from 2019 to 2020 in days of absence for mental ill health during March and April by region according to the cumulative prevalence of new Covid-19 sickness absence in those regions during March-April 2020. There was a clear correlation between the two measures (weighted Pearson correlation coefficient = 0.67)

Results were not materially altered in the sensitivity analyses when excluding 43,171 individuals with imputed data or changed job over the study period (data not shown).

## Discussion

This analysis of national data on sickness absence in NHS staff found that superimposed on a rising trend since the beginning of 2019, there was a >50% surge in new episodes of prolonged absence for mental ill health during March and April 2020. The increase, which coincided with the first two months of the Covid-19 epidemic in England, and largely receded in the following two months, was greatest in those aged >60 years, and among Asian and Black employees. Moreover, it varied by region, correlating with rates of Covid-19 sickness absence during the same period. However, it was not observed in doctors and dentists, and was lower than average in registered nurses.

The study used data that had been assembled prospectively in a standardised format on a cohort of nearly a million healthcare workers. Information on sex, age, staff group and region will have been highly reliable, and any misclassification between the broad categories of ethnicity should have been small. It is possible that there was some under-ascertainment of absences that lasted only for a day or two, but we would expect longer term sickness absence, and especially episodes with duration >28 days, to have been reliably recorded. Identification of Covid-19 as a reason for sickness absence will not have been completely accurate, especially in the early phase of the epidemic when diagnostic tests were not widely available. However, using data from two trusts, we have shown that Covid-19 sickness absence by our definition was associated with a substantially higher prevalence of positive results in later antibody tests (van der Plaat et al., 2021). Because of stigma, it is possible that some sickness absence attributable to mental ill health was inappropriately ascribed to other diagnostic categories, although that should have been less of a problem for longer duration absences, which would normally be supported by medical certification. Moreover, there seems no reason why impacts of stigma would have been lower in March and April of 2020 than both earlier and later.

Mental ill health is estimated to account for more than a quarter of sickness absence in NHS staff (ONS, 2021), the large majority of which is for common mental health disorders, including anxiety, depression and post-traumatic stress disorder (Boorman, 2009; ONS, 2021). Over the 18-month study period, almost 6% of employees experienced at least one episode of sickness absence for mental ill health that lasted longer than 28 days, and a further 7% had shorter absences for mental health problems. We did not have more specific diagnostic information, but it is likely that absence in most cases will have been for common mental health disorders, and only rarely for psychosis or organic psychiatric disease.

Whether or not mental ill health leads to absence from work will depend in part on the extent to which symptoms are tolerated, and thresholds for taking time off may be influenced by cultural norms within the workforce as well as by factors specific to the individual (Westerlund et al., 2004). It is possible that cultural differences account for the substantially lower risk of long-term absence for mental ill health that we observed in non-white as compared with White employees, even after allowance for other demographic and occupational characteristics. Little has been reported on the association between ethnicity and sickness absence. In England, belonging to a minority ethnic group is strongly associated with risk factors for mental ill health, as well as unemployment; lone parent status; lower social class; low social support and poverty. Evidence suggests that once these factors are taken into account, ethnic groups have a similar risk of common mental disorders (Brugha et al., 2004; Weich et al., 2004). Therefore after adjustment of other risk factors, we would expect to find sickness absence rates in minority ethnic groups, to reflect that of the white workers. The differences found in this study may be a reflection of the fact that mental ill health is considered to be highly stigmatising in some minority ethnic groups (Bignall, Jeraj, Helsby, & Butt, 2019) and this may lead to non-disclosure by the patient to the doctor who is issuing the fit note, and to the workplace. Our findings are not generalisable beyond healthcare workers in England and findings from other industries, may shed light on the interpreattaion of our findings.

Behavioural norms may also differ importantly between occupational groups. Thus, the lower rates of long-term absence for mental ill health among doctors, dentists and healthcare scientists than in most other staff groups may have been driven in part by a culture of presenteeism. After adjustment for other risk factors, risk in additional clinical services (a staff group that included care assistants and other less skilled work in support of patient care) was some four times that in medical and dental personnel.

The observed geographical differences in risk are striking, with rates of long-term absence for mental ill health increasing progressively with distance from London and the South-East. Furthermore, the same pattern was present when analysis was limited to specific staff groups, including doctors and dentists among whom risk in the North-West and North-East was two to three times that in London. Our regional results accord with longer term national data on sickness absence (all sickness absence, not just mental health) among NHS staff in England, which have following roughly the same pattern as our findings for the last decade (NHS Digital, 2021). Regional variation in NHS sickness absence rates for ‘stress’, available for 2017-2018, has similar distribution to that seen in our study (NHS Digital, 2018).

Temporal changes over the course of the study period in the burden of sickness absence from mental ill health appear to reflect two distinct phenomena, with a spike of absence episodes lasting >28 days in March and April of 2020 that was superimposed on a longer term trend of increasing rates, dating back to January 2019 or earlier. By comparing days lost from work in March and April of 2020 to the corresponding months in 2019, we were able to take out possible seasonal variation, and the overall year-on-year increase was substantial (73%).

That the percentage increase varied by region, in a way that correlated with cumulative prevalence of Covid-19 sickness absence during March and April 2020 (correlation coefficient = 0.67) supports the view that it was driven by stresses arising from the epidemic, either at work or domestically. It is, however, notable that that the impact differed by staff group, the relative increase being greatest in additional clinical services, estates and ancillary staff (which includednon-patient-facing roles such as gardeners, fitters and engineers) and healthcare scientists, with smaller percentage increases in registered nurses and allied health professionals (such as physiotherapists and occupational therapists), and a significant decline in medical and dental staff. It also varied by age (greater below age 35 years and particularly in those aged >60 years), and was somewhat greater in Black and Asian ethnic groups, although the latter may reflect, at least in part, that rates in those groups started from a lower baseline. It is likely that some workers in less skilled jobs came from poorer socioeconomic circumstances than those in professional roles, leading to greater pressures outside work as a consequence of the epidemic. And increased demands outside work (e.g. related to childcare during lockdown and financial worries) may have added to pressures on younger workers. The large increase in the oldest workers could have been influenced by worries about their greater vulnerability to Covid-19, and by their being closer to retirement. Another factor in the varying impact by staff group may have been differences in peer-group support and in the sense of bringing special skills to challenging and important work that was valued by others, even if physically and emotionally demanding.

While our findings confirm that the first wave of the Covid-19 epidemic had an important effect on disabling mental ill health in health care workers, they also put the scale of that impact in perspective. The increase in rates of new long-duration (>28 days) absence in March and April of 2020 was less than a doubling overall, and in almost all subgroups of workers. As such, it was less than the long-term variation that occurred between regions. Moreover, the surge of new long-duration absences had substantially declined by May to June, suggesting that there was no major reaction as immediate pressures at the height of the wave subsided.

## Ethical approval statement

Approval granted by the NHS Health Research Authority (reference 20/SC/0282). The study was registered at ISRCTN: 36352994

## Author contributions

All authors contributed to the planning, conduct, analyses and reporting of this manuscript as outlined below.

Diana van der Plaat (Statistician/research associate): was responsible for the statistical aspects of analysis and interpretation of the quantitative aspects of the study.

Rhiannon Edge (Lecturer in Population Health): was responsible for advising on study design, analysis and interpretation of results.

David Coggon (Emeritus Professor of Occupational and Environmental Medicine): was responsible for advising on methodological design, analysis and interpretation of results.

Martie van Tongeren (Professor of Occupational and Environmental Medicine): was responsible for advising on study design, analysis and interpretation of results.

Rupert Muiry (Research assistant): was responsible for reviewing the emerging literature and assisted in drafting the manuscript.

Vaughan Parsons (Research manager): was responsible for overseeing the set-up and delivery of the study, and facilitated data collection.

Paul Cullinan (Professor in Occupational and Environmental Respiratory Disease): was chief investigator with responsibility for advising on study design, analysis and interpretation of results. Had overall responsibility for the management and delivery of the study.

Ira Madan (Consultant Occupational Physician and Reader): was co-chief investigator with responsibility for advising on study design, analysis, interpretation of results and for drafting the manuscript.

## Data Sharing Statement

With permission, source data are available upon request from the NHS Electronic Staff Record (ESR) Warehouse (NHS England)

## Data Availability

Data Sharing Statement
With permission, source data are available upon request from the NHS Electronic Staff Record (ESR) Warehouse (NHS England)

## Acknowledgments

Sam Wright, Workforce Information Advisor, NHS Electronic Staff Record, and Mike Vickerman, Workforce Information and Analysis, DHSC. Dr Gavin Debrera (Public Health England) and Dr Kit Harling gave invaluable assistance in planning the study and sourcing the datasets.

We appreciate Lee Isidore, Manal Sadik and Victoria Thorpe from Guy’s and St Thomas NHS Foundation Trust for their helpful input into the interpretation of our findings.

We would also like to thank Cambridge University Hospitals NHS Foundation Trust (Dr Mark Ferris), Guys and St Thomas’s NHS Foundation Trust (Dr Ali Hashtroudi) and Bolton NHS Foundation Trust (Dr Martin Seed) for providing results from their staff antibody testing programmes.

