## Supplementary material for "IMPACT OF COVID-19 PANDEMIC ON SICKNESS ABSENCE FOR MENTAL ILL HEALTH IN NATIONAL HEALTH SERVICE STAFF": Tables and figures

### Table 1. Cumulative prevalence of new sickness absence for mental ill health at any time during 1 January 2019 to 30 June 2020 by demographic characteristics, staff group, region and duration of longest episode

| **Characteristic** | **Number at risk** | **At least one new episode of sickness absence for mental illness, but none with duration >28 days** | | | | **At least one new episode of sickness absence for mental illness with duration >28 days** | | | |
| --- | --- | --- | --- | --- | --- | --- | --- | --- | --- |
|  |  | **Cases** | **Cumulative prevalence (%)** | **^a^OR** | **95%CI** | **Cases** | **Cumulative prevalence (%)** | **^a^OR** | **95%CI** |
| **All subjects** | 959,356 | 65,104 | 6.8 | - | - | 54,421 | 5.7 | - | - |
| **Sex** |  |  |  |  |  |  |  |  |  |
| Female | 738,495 | 54,983 | 7.4 | *ref.* | *.* | 45,896 | 6.2 | *ref.* | *.* |
| Male | 220,861 | 10,121 | 4.6 | 0.72 | 0.70 - 0.73 | 8,525 | 3.9 | 0.71 | 0.70 - 0.73 |
| **Age (years)** |  |  |  |  |  |  |  |  |  |
| <25 | 26,217 | 2,819 | 10.8 | *ref.* | *.* | 1,125 | 4.3 | *ref.* | *.* |
| 25-29 | 90,398 | 7,982 | 8.8 | 0.92 | 0.88 - 0.97 | 4,164 | 4.6 | 1.24 | 1.16 - 1.32 |
| 30-34 | 114,249 | 8,750 | 7.7 | 0.83 | 0.79 - 0.87 | 6,136 | 5.4 | 1.50 | 1.40 - 1.60 |
| 35-39 | 110,193 | 7,586 | 6.9 | 0.75 | 0.71 - 0.78 | 6,707 | 6.1 | 1.69 | 1.58 - 1.81 |
| 40-44 | 120,865 | 7,777 | 6.4 | 0.72 | 0.68 - 0.75 | 6,913 | 5.7 | 1.64 | 1.54 - 1.75 |
| 45-49 | 133,797 | 8,581 | 6.4 | 0.70 | 0.67 - 0.73 | 7,901 | 5.9 | 1.65 | 1.55 - 1.76 |
| 50-54 | 141,791 | 9,259 | 6.5 | 0.68 | 0.65 - 0.71 | 8,788 | 6.2 | 1.61 | 1.51 - 1.72 |
| 55-60 | 141,925 | 8,613 | 6.1 | 0.60 | 0.58 - 0.63 | 8,479 | 6.0 | 1.47 | 1.38 - 1.57 |
| >60 | 79,921 | 3,737 | 4.7 | 0.45 | 0.43 - 0.48 | 4,208 | 5.3 | 1.25 | 1.17 - 1.34 |
| **Ethnicity** |  |  |  |  |  |  |  |  |  |
| White | 731,408 | 54,299 | 7.4 | *ref.* | *.* | 46,209 | 6.3 | *ref.* | . |
| South Asian | 66,881 | 2,565 | 3.8 | 0.63 | 0.60 - 0.65 | 2,064 | 3.1 | 0.62 | 0.59 - 0.65 |
| Other or unspecified Asian | 39,585 | 1,659 | 4.2 | 0.58 | 0.55 - 0.61 | 941 | 2.4 | 0.44 | 0.42 - 0.48 |
| Black | 56,494 | 2,707 | 4.8 | 0.71 | 0.68 - 0.73 | 2,050 | 3.6 | 0.67 | 0.64 - 0.70 |
| Mixed | 17,019 | 1,199 | 7.0 | 1.04 | 0.98 - 1.10 | 928 | 5.5 | 1.05 | 0.98 - 1.13 |
| Other | 13,434 | 625 | 4.7 | 0.77 | 0.71 - 0.84 | 406 | 3.0 | 0.62 | 0.56 - 0.68 |
| Unknown | 34,535 | 2,050 | 5.9 | 0.90 | 0.86 - 0.94 | 1,823 | 5.3 | 0.96 | 0.91 - 1.01 |
| **Staff group at 01-01-2019** |  |  |  |  |  |  |  |  |  |
| Administrative and clerical | 205,822 | 18,068 | 9.5 | *ref.* | *.* | 11,639 | 5.7 | *ref.* | . |
| Additional clinical services | 190,443 | 2,447 | 5.7 | 1.55 | 1.51 - 1.59 | 15,711 | 8.2 | 1.58 | 1.54 - 1.62 |
| Additional professional scientific and technical | 42,696 | 4,310 | 5.9 | 0.88 | 0.84 - 0.92 | 1,950 | 4.6 | 0.82 | 0.78 - 0.86 |
| Allied health professionals | 72,470 | 3,676 | 5.9 | 0.85 | 0.82 - 0.88 | 3,137 | 4.3 | 0.75 | 0.72 - 0.78 |
| Estates and ancillary | 62,104 | 1,043 | 4.7 | 1.04 | 1.00 - 1.08 | 3,667 | 5.9 | 1.09 | 1.05 - 1.13 |
| Healthcare scientists | 22,003 | 1,188 | 1.5 | 0.73 | 0.68 - 0.78 | 657 | 3.0 | 0.54 | 0.49 - 0.58 |
| Medical and dental | 80,267 | 21,124 | 7.6 | 0.26 | 0.24 - 0.27 | 1,256 | 1.6 | 0.32 | 0.30 - 0.34 |
| Nursing and midwifery registered | 279,619 | 100 | 5.0 | 1.17 | 1.14 - 1.19 | 16,241 | 5.8 | 1.05 | 1.02 - 1.08 |
| Students | 1,990 | 133 | 6.8 | 0.60 | 0.49 - 0.74 | 35 | 1.8 | 0.31 | 0.22 - 0.44 |
| Multiple and unknown | 1,942 | 5,537 | 4.2 | 1.05 | 0.88 - 1.25 | 128 | 6.6 | 1.17 | 0.98 - 1.40 |
| **Region** |  |  |  |  |  |  |  |  |  |
| London | 133,378 | 5,537 | 4.2 | *ref.* | *.* | 3,893 | 2.9 | *ref.* | *.* |
| South East | 131,568 | 9,224 | 7.0 | 1.55 | 1.5 - 1.61 | 5,336 | 4.1 | 1.23 | 1.18 - 1.29 |
| East of England | 82,547 | 5,994 | 7.3 | 1.59 | 1.53 - 1.65 | 3,960 | 4.8 | 1.45 | 1.38 - 1.52 |
| South West | 97,420 | 7,308 | 7.5 | 1.61 | 1.55 - 1.67 | 4,926 | 5.1 | 1.47 | 1.41 - 1.54 |
| East Midlands | 66,525 | 5,216 | 7.8 | 1.72 | 1.65 - 1.79 | 4,131 | 6.2 | 1.86 | 1.78 - 1.95 |
| Yorkshire and the Humber | 121,775 | 9,278 | 7.6 | 1.64 | 1.58 - 1.70 | 8,110 | 6.7 | 1.95 | 1.87 - 2.03 |
| West Midlands | 110,474 | 7,493 | 6.8 | 1.50 | 1.44 - 1.55 | 6,964 | 6.3 | 1.91 | 1.83 - 1.99 |
| North East | 55,266 | 3,904 | 7.1 | 1.51 | 1.44 - 1.57 | 4,493 | 8.1 | 2.34 | 2.23 - 2.45 |
| North West | 160,403 | 11,150 | 7.0 | 1.55 | 1.50 - 1.60 | 12,608 | 7.9 | 2.36 | 2.27 - 2.45 |

^a^Odds ratio and 95% confidence interval from a multiple logistic regression analysis that included all of the variables for which results are presented. The reference was individuals with no absence for mental ill health.

### Figure 1. Sickness absence for mental ill health during 1 January 2019 to 31 June 2020: total days lost and numbers of new episodes by time period

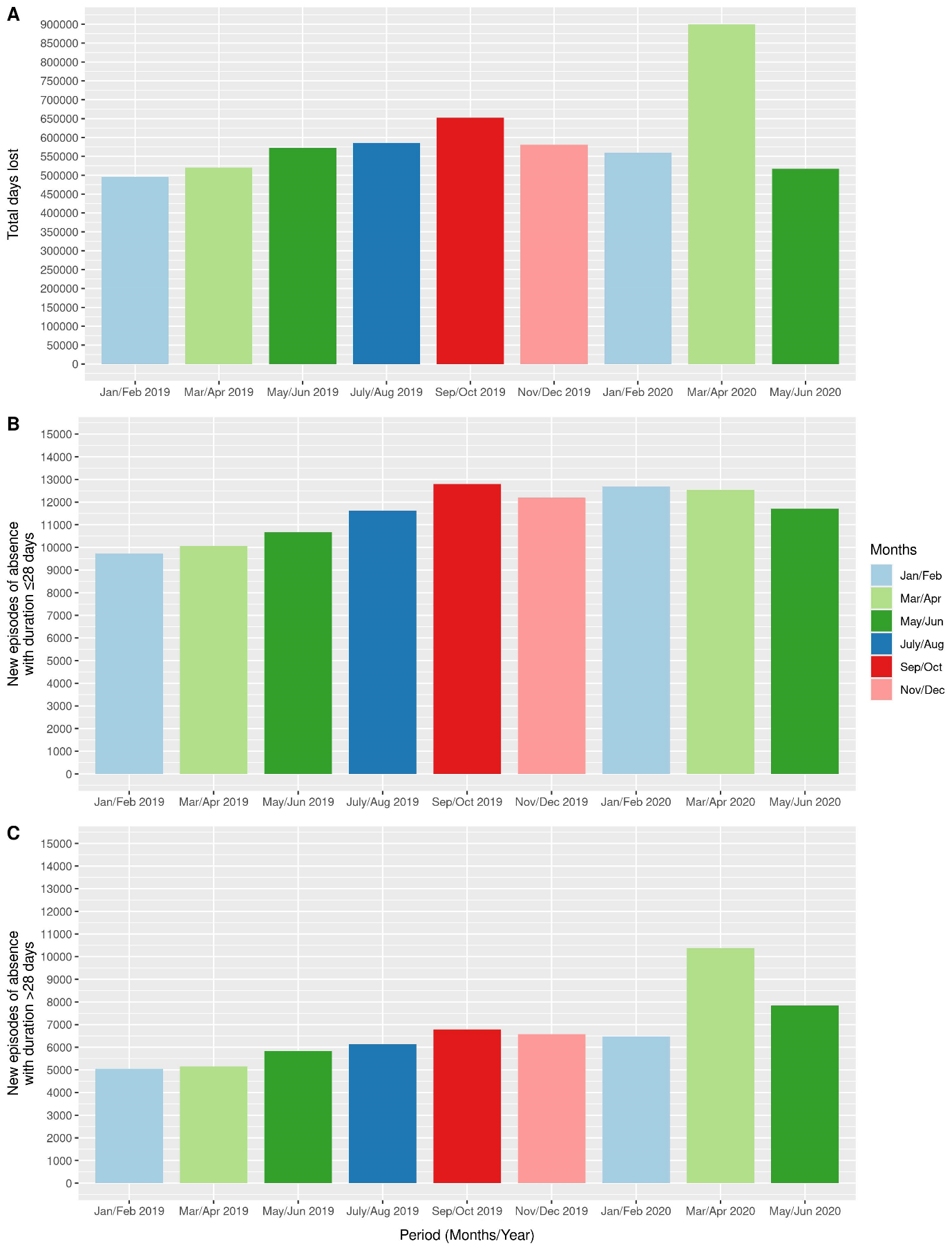

### Table 2. Total days lost through sickness absence for mental ill health during March-April 2019 and March-April 2020 according to demographic characteristics, staff group and region

| **Characteristic** | **Total days lost through sickness absence for mental illness** | | **Percentage change from 2019 to 2020 (95%CI)** | |
| --- | --- | --- | --- | --- |
|  | **March-April 2019** | **March-April 2020** |  |  |
| All subjects | 519,807 | 899,730 | 73.1 | (72.5 to 73.7) |
| **Sex** |  |  |  |  |
| Female | 435,003 | 774,499 | 78.0 | (77.4 to 78.7) |
| Male | 84,804 | 125,231 | 47.7 | (46.4 to 49.0) |
| **Age (years)** |  |  |  |  |
| <25 | 9,530 | 19,440 | 104 | (99 to 109) |
| 25-29 | 32,507 | 72,733 | 124 | (121 to 127) |
| 30-34 | 52,255 | 105,567 | 102 | (100 to 104) |
| 35-39 | 66,489 | 106,070 | 59.5 | (58.0 to 61.1) |
| 40-44 | 69,594 | 106,396 | 52.9 | (51.4 to 54.3) |
| 45-49 | 79,996 | 116,861 | 46.1 | (44.8 to 47.4) |
| 50-54 | 89,712 | 120,680 | 34.5 | (33.4 to 35.7) |
| 55-60 | 84,928 | 138,310 | 62.9 | (61.5 to 64.3) |
| >60 | 34,796 | 113,673 | 227 | (223 to 231) |
| **Ethnicity** |  |  |  |  |
| White | 438,624 | 742,614 | 69.3 | (68.7 to 69.9) |
| South Asian | 20,383 | 42,525 | 109 | (105 to 112) |
| Other or unspecified Asian | 8,522 | 20,147 | 136 | (130 to 142) |
| Black | 17,014 | 39,376 | 131 | (127 to 136) |
| Mixed | 9,756 | 15,251 | 56.3 | (52.4 to 60.3) |
| **Staff group at 01-01-2019** |  |  |  |  |
| Administrative and clerical | 112,278 | 195,426 | 74.1 | (72.8 to 75.3) |
| Additional clinical services | 142,018 | 281,286 | 98.1 | (96.8 to 99.3) |
| Additional professional scientific and technical | 21,172 | 29,251 | 38.2 | (35.7 to 40.6) |
| Allied health professionals | 27,941 | 43,135 | 54.4 | (52.1 to 56.7) |
| Estates and ancillary | 30,953 | 76,603 | 148 | (144 to 151) |
| Healthcare scientists | 6,128 | 11,112 | 81.3 | (75.8 to 87.1) |
| Medical and dental | 19,688 | 17,188 | -12.7 | (-14.5 to -10.9) |
| Nursing and midwifery registered | 158,011 | 243,015 | 53.8 | (52.8 to 54.8) |
| **Region** |  |  |  |  |
| London | 32,617 | 72,373 | 122 | (119 to 125) |
| South East | 50,644 | 86,168 | 70.1 | (68.3 to 72.0) |
| East of England | 36,564 | 71,501 | 95.6 | (93.1 to 98.0) |
| South West | 51,527 | 75,118 | 45.8 | (44.2 to 47.4) |
| East Midlands | 42,607 | 61,241 | 43.7 | (42.0 to 45.5) |
| Yorkshire and the Humber | 80,085 | 138,655 | 73.1 | (71.6 to 74.6) |
| West Midlands | 61,109 | 113,885 | 86.4 | (84.5 to 88.2) |
| North East | 42,752 | 72,454 | 69.5 | (67.5 to 71.5) |
| North West | 121,902 | 208,335 | 70.9 | (69.7 to 72.1) |

### Figure 2. Percentage change from 2019 to 2020 in days of absence for mental ill health during March and April by region according to the cumulative prevalence of new Covid-19 sickness absence during March and April 2020

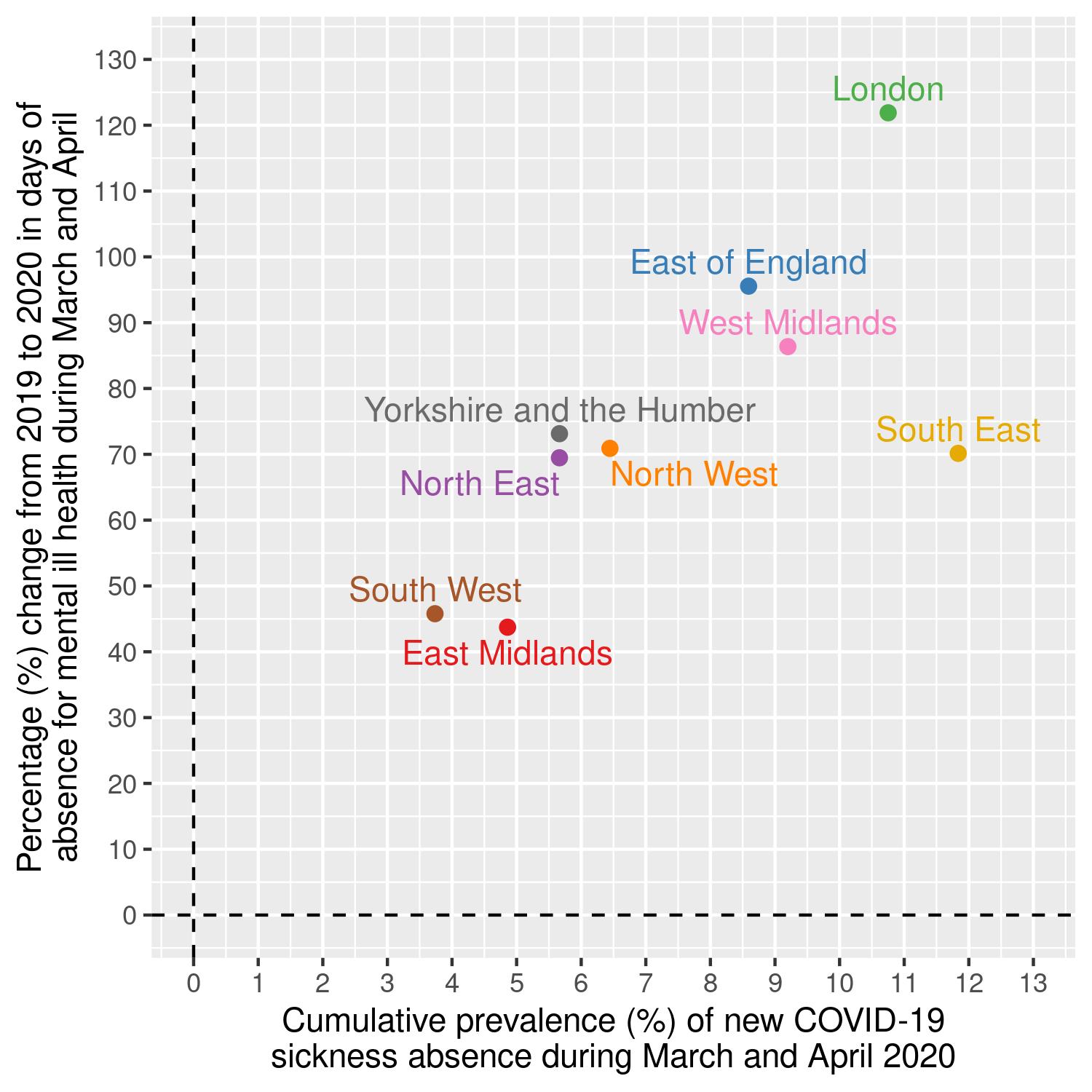

### Supplementary Table S1. Cumulative prevalence of new long-term sickness absence for back problems and other musculoskeletal disorders during 1 January 2019 to 30 June 2020 by staff group and region

| **Characteristic** | **^a^Cases** | **Cumulative prevalence (%)** | **^b^OR** | **95%CI** |
| --- | --- | --- | --- | --- |
| **All subjects** | 18,931 | 2.0 |  |  |
| **Staff group at 01-01-2019** |  |  |  |  |
| Administrative and clerical | 3,037 | 1.5 | *ref.* | *.* |
| Additional clinical services | 5,927 | 3.1 | 2.59 | 2.48 - 2.71 |
| Additional professional scientific and technical | 581 | 1.4 | 1.08 | 0.99 - 1.18 |
| Allied health professionals | 1,105 | 1.5 | 1.29 | 1.21 - 1.39 |
| Estates and ancillary | 2,205 | 3.6 | 2.43 | 2.30 - 2.58 |
| Healthcare scientists | 178 | 0.8 | 0.64 | 0.55 - 0.74 |
| Medical and dental | 388 | 0.5 | 0.35 | 0.31 - 0.38 |
| Nursing and midwifery registered | 5,446 | 1.9 | 1.56 | 1.50 - 1.64 |
| Students | 18 | 0.9 | 1.07 | 0.67 - 1.71 |
| Multiple and unknown | 46 | 2.4 | 1.81 | 1.34 - 2.43 |
| **Region** |  |  |  |  |
| London | 1,812 | 1.4 | *ref.* | *.* |
| South East | 2,123 | 1.6 | 1.08 | 1.01 - 1.15 |
| East of England | 1,420 | 1.7 | 1.14 | 1.06 - 1.23 |
| South West | 1,506 | 1.5 | 0.98 | 0.91 - 1.05 |
| East Midlands | 1,342 | 2.0 | 1.23 | 1.15 - 1.33 |
| Yorkshire and the Humber | 2,785 | 2.3 | 1.38 | 1.30 - 1.48 |
| West Midlands | 2,415 | 2.2 | 1.36 | 1.28 - 1.45 |
| North East | 1,533 | 2.8 | 1.59 | 1.48 - 1.71 |
| North West | 3,995 | 2.5 | 1.53 | 1.44 - 1.62 |

^a^Individuals with at least one new episode of sickness absence, either for back problems or for other musculoskeletal disorders, that lasted for >28 days

^b^Odds ratios (with 95% confidence intervals) from a single logistic regression model that also included sex, age and ethnicity, all of which were classified as in Table 1.
